# Modeling the Stability of Severe Acute Respiratory Syndrome Coronavirus 2 (SARS-CoV-2) on Skin, Currency, and Clothing

**DOI:** 10.1101/2020.07.01.20144253

**Authors:** David E. Harbourt, Andrew D. Haddow, Ashley E. Piper, Holly Bloomfield, Brian J. Kearney, David Fetterer, Kathleen Gibson, Timothy Minogue

**Affiliations:** Biosafety Division, United States Army Medical Research Institute of Infectious Diseases, 1425 Porter Street, Ft. Detrick Maryland, 21702; General Dynamics Health Solutions in support of USAMRIID, 1425 Porter Street, Ft. Detrick, Maryland, 21702; Oak Ridge Institute of Science and Education, 1425 Porter Street, Ft. Detrick, Maryland, 21702; Core Laboratory Services Directorate, United States Army Medical Research Institute of Infectious Diseases, 1425 Porter Street, Ft. Detrick Maryland, 21702; ICON Global Public Health Solutions, 1425 Porter Street, Ft. Detrick, Maryland, 21702; Diagnostic Systems Division, United States Army Medical Research Institute of Infectious Diseases, 1425 Porter Street, Ft. Detrick Maryland, 21702

## Abstract

A new coronavirus (SARS-CoV-2) emerged in the winter of 2019 in Wuhan, China, and rapidly spread around the world. The extent and efficiency of SARS-CoV-2 pandemic is far greater than previous coronaviruses that emerged in the 21^st^ Century. Here, we modeled stability of SARS-CoV-2 on skin, paper currency, and clothing to determine if these surfaces may factor in the fomite transmission dynamics of SARS-CoV-2. Skin, currency, and clothing samples were exposed to SARS-CoV-2 under laboratory conditions and incubated at three different temperatures (4°C± 2°C, 22°C± 2°C, and 37°C ± 2°C). Stability was evaluated at 0 hours (h), 4 h, 8 h, 24 h, 72 h, 96 h, 7 days, and 14 days post-exposure. SARS-CoV-2 was shown to be stable on skin through the duration of the experiment at 4°C (14 days). Virus remained stable on skin for at least 96 h at 22°C and for at least 8h at 37°C. There were minimal differences between the tested currency samples. The virus remained stable on the $1 U.S.A. Bank Note for at least 96 h at 4°C while viable virus was not detected on the $20 U.S.A. Bank Note samples beyond 72 h. The virus remained stable on both Bank Notes for at least 8 h at 22°C and 4 h at 37°C. Clothing samples were similar in stability to the currency with the virus being detected for at least 96 h at 4°C and at least 4 h at 22°C. No viable virus was detected on clothing samples at 37°C after initial exposure. This study confirms the inverse relationship between virus stability and temperature. Furthermore, virus stability on skin demonstrates the need for continued hand hygiene practices to minimize fomite transmission both in the general population as well as workplaces where close contact is common.

## Background

The emergence of SARS-CoV-2 represents the third major outbreak of a new human coronavirus disease over the past twenty years. This novel coronavirus (SARS-CoV-2) that initially emerged in Wuhan, China in late 2019 resulted in a global pandemic that, as of April 22, 2020, has officially resulted in more than 2.5 million cases and 175,000 deaths (1–3). The rapid and extensive spread of the virus could be indicative of both aerosol and fomite transmission which has been seen in previous coronavirus outbreaks (4). Previous studies have shown that SARS-CoV-2 is stable at room temperature on stainless steel for approximately 24 hours (h) and on cardboard for up to three days (5–6). Additional studies have shown an inverse relationship between surface temperature and stability which is consistent with previous reporting on stability of human coronaviruses (5).

While some aspects of SARS-CoV-2 have been reported, there have been only limited investigations into stability on skin specimens and paper currency (7). Despite limited evidence, some countries have taken measures to limit the spread of the virus by either burning or disinfecting paper currency or discouraging the use of cash during transactions (8). It would be expected that the stability of SARS-CoV-2 on currency would be similar to cardboard given that both surfaces are porous, but the effect of ink and toner on the virus remains unknown (6).

Furthermore, it was not known how long SARS-CoV-2 could remain viable on human or animal skin as no similar studies had been performed to date. Handwashing and hand hygiene have been a key part of mitigation efforts, but fomite transmission likely remains a contributing factor to the speed and extent of the pandemic (9–11). Herein, we model the stability of SARS-CoV-2 across animal skin, paper currency, and clothing.

## Methods and Materials

### Virus isolate

We utilized the USA-WA1/2020 strain of SARS-CoV-2 isolated from a human patient in Washington State, USA, in January 2020 (GenBank accession no. MN985325.1). This isolate was selected due to its use in related studies (6). A working virus stock was prepared by adding virus to Grivet (*Chlorocebus aethiops*) Vero 76 kidney cells (ATCC, Manassas, VA; #CRL-1587) at a multiplicity of infection (MOI) of 0.01. Cells were incubated for 1 h for virus adsorption and maintained in Eagles Minimal Essential Media (EMEM) with 10% fetal bovine serum at 5% CO_2_. The cell supernatant was harvested 50 h post-inoculation. The supernatant was clarified at 10,000 x *g* for 10 minutes at 4°C and stored at −70°C until use.

### Virus surface stability

We evaluated the surface stability of SARS-CoV-2 on four common surfaces (approx. 6.3 mm^2^). These included: swine skin (*Sus scrofa*) with the hair removed (acquired from a local butcher); uncirculated United States of America $1 and $20 Federal Reserve notes comprised of 25% linen and 75% cotton with red and blue security fibers (United States Secret Service, Washington, DC, USA); and unused scrub fabric consisting of 35% cotton and 65% polyester (Labforce, Swedesboro, NJ, USA). Using a pipette, we deposited 50 µL of virus onto the surface of each material in triplicate. Individual groups of samples were incubated for 0 h, 4 h, 8 h, 24 h, 72 h, 96 h, 7 days, and 14 days post-exposure across three temperatures; 4°C± 2°C, 22°C± 2°C, and 37°C ± 2°C at a relative humidity of 40-50%. Following incubation, samples were transferred to 2-mL CryoSure tubes (Caplugs Evergreen, Buffalo, NY, USA) containing 1 mL of media minimum essential media (MEM: Corning, catalog 10-010-CM), supplemented with 4.0 µg/mL gentamicin (GIBCO, Carlsbad, CA, USA), 2% Penicillin (Sigma-Aldrich, St. Louis, MO, USA), 2% streptomycin (Sigma-Aldrich), and 2.5 mg/mL of amphotericin B (Sigma-Aldrich, St. Louis, MO, USA) using featherweight forceps (BioQuip Products, Inc, Rancho Domingo, CA, USA) to reduce the potential for sample damage. Forceps were disinfected using 5% Microchem Plus™ followed by 70% ETOH between samples. Samples were stored at −80°C prior to virus quantification.

### Detection and quantification of infectious virus

Confluent cultures (90-95% confluency) of ATCC Vero 76 cells in 6-well plates were utilized for all assays. Samples were thawed at ambient temperature and diluted by performing a series of 1:10 dilutions in MEM + 5% Heat Inactivated (HI) FBS + 2% penicillin, 2% streptomycin + 0.5% fungizone (MEM Complete). All samples were assayed as undiluted and up to three additional ten-fold dilutions. Media was removed from plates and cells were infected with 100 µL of sample in triplicate. Cells were incubated at 37°C and 5% CO_2_ for one hour, with rocking approximately every 15 minutes. Following incubation, media-agarose overlay (2mL of a 1:1 mixture of 1.0% agarose and 2X EBME + 10% HI FBS + 2% penicillin, 2% streptomycin + 1% fungizone (2X EBME Complete)) was added to each well. Once overlay solidified, plates were incubated at 37°C and 5% CO_2_ for 48 h ± 4 h. Following incubation, a second media agarose overlay containing 4% neutral red in a 1:1 mixture of 1.0% agarose and 2X EBME complete was added to each well. Once overlay solidified, plates were incubated at 37°C and 5% CO_2_ overnight. Following incubation, plaques were counted and the virus yield (plaque forming unit, PFU/mL) for each sample calculated, with a lower limit of quantification (LLOQ) of 2.0 log_10_ PFU/mL. The limit of detection (LOD) for virus isolation attempts was 0.1 log_10_ PFU/mL.

### Statistical analyses

The half-life was estimated by fitting, to each trial, a log-linear Poisson regression model of the form

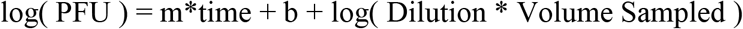

such that log(Dilution * Volume Sampled) was the offset. The half-life was estimated as –log(2)/m. This analysis was implemented in SAS/PROC GENMOD. The log of the half-lives so estimated were entered into a two-way ANOVA, as implemented in SAS/PROC MIXED. The mixed model procedure was used to allow for a heterogeneous variance structure (11). Denominator degrees of freedom were estimated by Satterthwaite’s method, and LS-mean differences between temperatures and surfaces were evaluated. For the purpose of comparing half-lives, one outlier was removed from the clothing surface at 22°C, pursuant to a Dixon gap test applied to the log transformed half-life. Analysis was implemented in SAS version 9.4 (SAS Institute, Cary, NC), with the exception of the gap test, which was implemented in R (12) package outliers version 0.14. No adjustment for multiplicity has been applied to the reported p-values.

## Results

SARS-CoV-2 remained stable on skin at 4°C for the duration of the experiment (Fig 1 and Fig 2). The virus exhibited similar initial decay profiles at 4°C across all surfaces. Initially, the virus exhibited a loss of 1-2 log_10_ PFU in the first 8 h across all surfaces. However, after 8 h, the virus appeared to stabilize to varying degrees for the remainder of the experiment. Clothing and $1 U.S.A. Bank Note samples remained viable until 96 h with $20 U.S.A. Bank Note samples remaining detectable for 7 days. While the virus continued to lose the remaining 2-3 log_10_ PFU over the remaining time points of the experiment, the decay rate at 4°C was slower on skin than any other tested surface.

**Fig 1.**
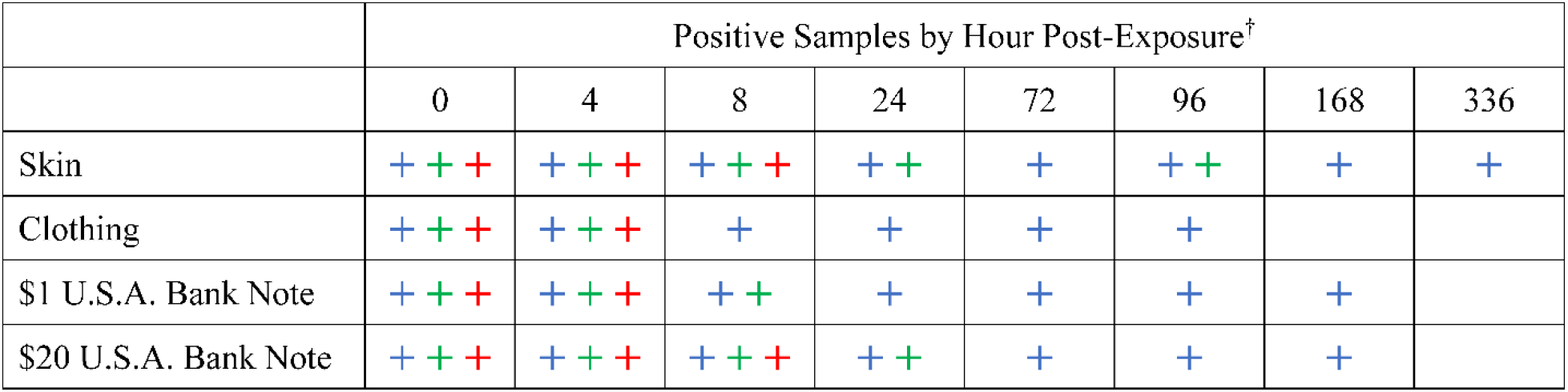
Recovery of infectious virus. Limit of detection was one plaque forming unit ^†^Not tested: Skin (22°C at 336 h post-exposure, 37°C at 168 and 336 h post-exposure); and Cloth, $1 U.S.A. Bank Note, and $20 U.S.A. Bank Note (22°C and 37°C, at 168 h and 336 h post-exposure) Positive samples at 4°C are represented by a blue + Positive samples at 22°C are represented by a green + Positive samples at 37°C are represented by a red +

**Fig 2.**
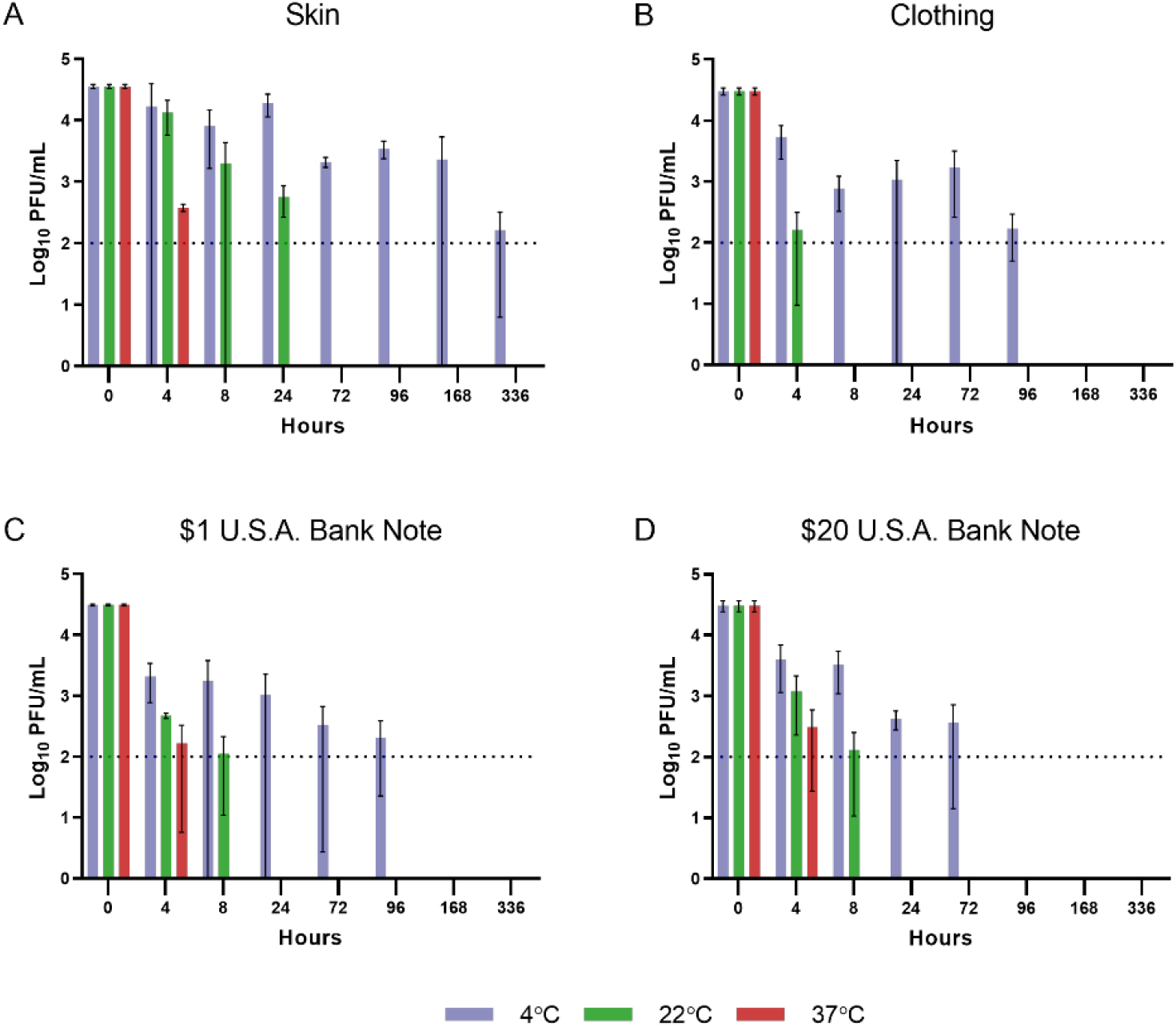
Quantification of infectious virus. Viral concentrations were determined as described in METHODS. Viral concentrations are expressed as mean ± SD log_10_ PFU/mL. ^†^Not tested: Skin (22°C at 336 h post-exposure, 37°C at 168 h and 336 h post-exposure); and Cloth, $1 U.S.A. Bank Note, and $20 U.S.A. Bank Note (22°C and 37°C, at 168 h and 336 h post-exposure) *Lower limit of quantification (LLOQ) of 2.0 log_10_ PFU/mL

At 22°C, the virus appeared to lose approximately 2-3 log_10_ PFU within the first 8 h across all surfaces (Fig 2). Virus was not isolated from the $1 U.S.A. Bank Note or clothing beyond 8 h at room temperature. Viable virus was detected on the $20 U.S.A. Bank Note at 24 h but all subsequent samples were negative for infectious virus (Fig 1 and Fig 2). Approximately 3 log_10_ PFU of viable virus was detectable on the skin samples at 24 h at 22°C. Virus was isolated at 96 h but all other tested samples were below the LOD.

SARS-CoV-2 remained viable at 37°C on skin samples for up to 8 h (Fig 1 and Fig 2). No viable, infectious virus was detected on clothing samples after four hours at 37°C (Fig 2). There were minor differences in stability at 37°C between the $1 U.S.A. Bank Note and the $20 U.S.A. Bank Note, but those differences were not statistically significant. There was a discernable difference in virus stability across each of the temperatures with the 4°C conditions being the most hospitable conditions for virus stability even at 50% RH.

Statistical analysis indicated that skin samples had the longest half-life at each of the tested temperatures compared to the other surfaces (Table 1). Significant differences in virus stability were noted when each temperature condition was compared (Fig S1). Significant differences were observed in virus stability between the skin samples and all other tested surfaces (Fig S2). No other significant differences were observed during direct comparisons of the other tested surfaces.

**Table 1:** Estimates of Geometric Mean Half-Life by Temperature and Surface

|  | Temperature |  |  |
| --- | --- | --- | --- |
|  | 4°C | 22°C | 37°C |
| Skin | 46.8 ( 65.7 , 33.3 ) | 3.5 ( 14.6 , 0.9 ) | 0.6 ( 0.9 , 0.4 ) |
| Clothing | 33.7 ( 87.6 , 12.9 ) | 1.0 ( 1.6 , 0.6 ) | 0.2 ( 0.7 , 0.1 ) |
| \$1 U.S.A. Bank Note | 33.2 ( 77.1 , 14.3 ) | 1.3 ( 5.8 , 0.3 ) | 0.4 ( 2.8 , 0.1 ) |
| \$20 U.S.A Bank Note | 15.9 ( 79.1 , 3.2 ) | 1.1 ( 1.8 , 0.7 ) | 0.6 ( 1.3 , 0.3 ) |

Values indicate geometric mean (95%CL) of the half-life, in hours.

## Discussion

While the outbreak of SARS-CoV in 2002-2003 resulted in approximately 8000 confirmed cases and the 2012 MERS-CoV outbreak has culminated in less than 2500 cases, the current SARS-CoV-2 pandemic has resulted in over 2.5 million cases as of this writing and has spread to nearly every country on Earth (1–3). There are likely a number of reasons for the rapid emergence of SARS-CoV-2. Possibilities for enhanced spread include a different human infection case profile, virus transmission through asymptomatic carriers, or potentially enhanced fomite or aerosol transmission capability (2–4, 6, 7). Previous reports indicate that SARS-CoV-2 is more stable on cardboard and plastic compared to SARS-CoV, but aerosol decay rates are similar under laboratory conditions (6).

We have found that the skin samples were most hospitable for SARS-CoV-2, especially under refrigerated conditions. There are noticeable differences in virus decay rates at increasing temperatures which aligns with previous literature (13). Similar stability profiles were observed in both the currency and clothing samples. All three samples retained viable virus out to at least 96 h at 4°C with no recoverable virus beyond 8 h at 22°C in any of the three surfaces. There were small differences in viral concentration between the $1 U.S.A. Bank Note and $20 U.S.A. Bank Note samples, but they were not statistically significant. It is possible that differences in ink type, concentration, or both, affected virus stability and could warrant further study.

This is the first report to our knowledge modeling the stability of SARS-CoV-2 on skin. We note that even at 22°C, SARS-CoV-2 remained infectious on skin samples for 96 h post-exposure. While we understand real-world conditions cannot be replicated in the laboratory setting, this observation indicates the potential for fomite transmission in indoor environments in the absence of good hand hygiene practices, given that even trained medical students have been observed touching their faces approximately 23 times per hour (14). Furthermore, while we did use swine skin samples as a substitute for human skin, swine skin has enough similarities to human skin that it has been used for human allograft transplantation especially in burn victims (15). Based on this, it is expected that if this experiment were ever replicated using human skin samples, it would generate similar results.

We report the results of SARS-CoV-2 stability on animal skin as a series of outbreaks have been reported in the United States meat packing industry. Since most meat packing and processing procedures are carried out between 4-8°C, it is likely that any viral shedding from either symptomatic or asymptomatic workers in the absence of appropriate PPE would remain viable for an extended period of time on the surface of meat products or other surfaces (16–17). However, even with extensive cleaning, transmission could still occur in the presence of asymptomatic, undiagnosed workers due to both the enhanced stability of the virus and the high viral loads even asymptomatic cases maintain in the nasal passages (18). Without an extensive testing and contact tracing program, transmission around meat packing plants will likely continue to be an issue until herd immunity is reached either by infection or through the administration of an efficacious vaccine.

It is important to note the limitations of this study when analyzing the results. The variability in the decay curves between earlier studies and our results could have been due to differences in exposure doses (4, 6, 7). In addition, inherent variability with a study where virus is recovered from a tested surface can potentially confound some of the results, particularly at virus concentrations close to the limit of detection (LOD) or lower limit of quantification (LLOQ) (Fig. 1).

The results in this study demonstrate the continued inverse relationship between virus stability and temperature which is seen both in the laboratory and in the field when evaluating different transmission rates of SARS-CoV-2 in different parts of the world (19). While both fomite and aerosol transmission could be significant factors, due to the stability of SARS-CoV-2 on skin there is a continued need to reinforce proper hand hygiene practices and social distancing guidelines to minimize ongoing transmission potential.

## Data Availability

All data samples referred to in the manuscript are either included in the manuscript or supplementary files.

## Acknowledgements

The authors are indebted to Mr. Brian Sauerbry who assisted with material procurement, Dr. Pamela Glass for technical assistance and Ms. Sarah Norris for assistance with the statistical analysis. The views expressed in this article are those of the authors and do not reflect the official policy or position of the U.S. Department of Defense or the U.S. Army.

## Supplementary Figures

Supplementary Figure 1. Geometric mean ratio of half-lives, averaged across all three temperatures.

Supplementary Figure 2. Geometric mean ratio of half-lives, averaged across all four surfaces.

